# How do innovative occluding glasses compare with handheld spoon shaped occluders in children’s visual acuity test? A diagnostic agreement study

**DOI:** 10.64898/2026.09.15.26363094

**Authors:** Lu Tian, Shu Yang, Qing Zhou, Yuhan Song, Yuting Kan, Meizhen Zhao, Yan Jiang

## Abstract

**Objective:** To explore how a novel set of occluding glasses compare with handheld spoon shaped occluders in young children in terms of visual acuity test results, and how children felt about the two tools.

**Setting:** Outpatient clinic of the ophthalmology department in a university teaching hospital in Wuhan, China.

**Participants:** 210 children aged 3-9 yeas visiting the clinic for VAT from June 1 through July 30,2024 were conveniently selected as participants, among whom, 10 were excluded as their parents did not sign the informed consent paper.

**Study design:** This diagnostic agreement study compared two types of eye occluders using a cross-over design. The 200 children were divided into two equal batches: the first batch (case no.1-100) received VAT using handheld spoon-shaped eye occluders followed by occluding glasses, while the second batch (case no.101-200) received the interventions in the reverse order.

**Outcome measures:** Paired VAT scores generated from the study were compared with the Bland-Altman analysis. All children were asked of their acceptance regarding the two eye-covering tools upon completion of the 2nd VAT.

**Results:** The VAT score measured with occluding glasses were closely comparable with those measured with the handheld spoon-shaped eye occluders, with interclass correlation coefficient (ICC) of 0.989 and 0.989 for left eyes and right eyes, respectively(P<0.001). The mean difference of VAT score measured by the 2 tools was 0.007 for right eyes versus 0.005 for left eyes, and the 95% limits of agreement (LoA) ranged from −0.07 to 0.08 for right eyes versus 0.07 to 0.08 for left eyes, with 0.5 % versus 1 % of the measurements outside the 95% LoA. 61%(122/200) of the children preferred the appearance of the occluding glasses and considered it “more attractive than the handheld one”. However, respectively 90.0% and 93.0% of the children favored the hand-held occluder in terms of convenience and comfort.

**Conclusions:** The VAT measurements obtained with the two types of eye occluders showed excellent agreement, justifying the use of the occluding glasses as a promising tool for VAT in young children. Yet,refinement of the novel product is needed to enhance convenience and comfort.

## 1. INTRODUCTION

Myopia has become a significant public health concern, with projections indicating that its prevalence could reach 50% globally by 2050[1]. In China, rapid societal and economic development has led to lifestyle changes and increased educational pressures, resulting in a high incidence of myopia characterized by early onset, severe progression, and alarming rates of growth, particularly among children and adolescents[2]. According to a meta-analysis published in 2025, the prevalence of myopia among children and adolescents in China was approximately 58% (pooled weighted average effect size: 0.58, 95% CI: 0.54–0.62) [3].The ramifications of childhood myopia are profound, affecting lifelong development and quality of life. Research has demonstrated that more severe myopia resulted in a larger quality of life(QOL) decrease in both children and parents[4]. Moreover, childhood myopia not only impedes daily activities but also compromises self-esteem, which may in turn lead to heightened fear and increased exposure to stressful events or situations[5, 6]. Furthermore, if myopia is not corrected promptly or is improperly managed, it can progress to high myopia in adulthood, significantly increasing the risk of various blinding eye conditions, including primary open-angle glaucoma, retinal detachment and macular degeneration[7].Scholars have urged that focus on vision prevention and control should move forward to preschool children[8]. In response to this urgent call, the National Disease Control and Prevention Administration issued a guideline, explicitly proposes “shifting the gateway of vision health management forward,” designating “infancy, preschool, and the lower grades of primary school” as critical stages for myopia prevention and control[9].

Visual acuity test(VAT) is used to test an individual’s ability to distinguish different optotypes (stylized letters or symbols) at a standard distance [10]. It allows for quick and accurate differentiation of suspected myopia from children with normal vision. In 2021, the General Office of the Ministry of Education of China issued guidance mandating annual dynamic monitoring of the vision of children and adolescents nationwide, aiming for a coverage rate of over 90% for eye examinations among children[11]. Currently, various types of visual acuity charts are available for these examinations, including the Snellen, Landolt chart, as well as the tumbling E eye chart that prevails in most eye clinic in China. Regardless of which ever chart used, the examination requires the individual to cover one eye with an item that completely blocks the vision of the covered eye. In China, the most commonly used item for covering the eyes during VAT is handheld spoons. This tool may present some limitations, as children might apply pressure to or try to steal a glance from the covered eye, which could affect the accuracy of the vision test[12].

Based on the above-mentioned limitations, we developed a novel frame-shamed eye covering tool which was tailored to children’s mentality, and secured a national patent in China(ZL2023 2 1977842.3). Yet,how it compares with the routinely used handheld spoon-shaped eye occluders in terms of VAT scores, remains unclear. Therefore, we conducted a diagnostic agreement study to compare the new tool with traditional handheld spoon-shaped eye occluders, striving to determine the consistency of the results measured by the two tools and their acceptability in young children. Since the nasal bridge of the occluding glasses is a fixed-width component that cannot be adjusted to accommodate different facial widths in children, this study included only children aged 3 to 9 years.

## 2. Method

### 2.1 Study design

This diagnostic agreement study compared two types of eye occluders using a cross-over design. The 200 cases were divided into two equal batches: the first batch (case no.1-100) received VAT using handheld spoon-shaped occluders followed by occluding glasses, while the second (case no.101-200) received the tests in the reverse order. We then compared the two tools in terms of the visual acuity scores and children’s feedback regarding the tools’ appearance, ease of use, and comfort. The flowchart of the study was shown in Fig 1. This study was approved by the Medical Ethics Committee of Tongji Hospital, Tongji Medical College, Huazhong University of Science and Technology (TJ-IRB202403059).

**Fig 1.**
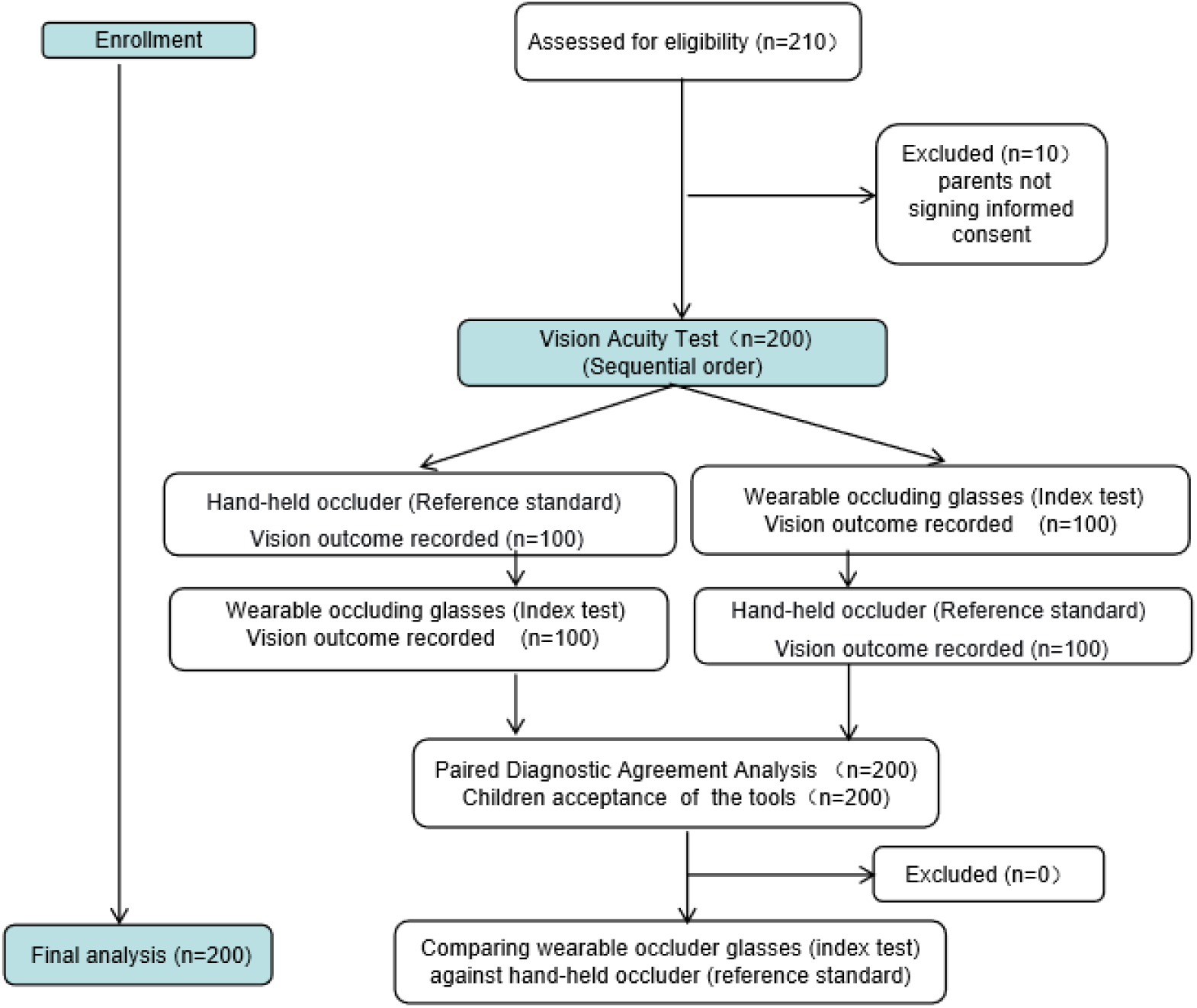
The flowchart of the study

### 2.2 Participants

We conducted this study at the ophthalmology outpatient clinic of a tertiary teaching hospital in Wuhan, Hubei Province, from June to July 2024. Considering that preschool and early school age is a critical period for children’s visual development, and the relatively high number of children aged 3 to 9 years attending the clinic [13], we included participants that met the following criteria: ① children aged 3 to 9 years; and ②children whose parents agreed to participate. We excluded: ① children who met World Health Organization’s diagnostic criteria for moderate or severe visual impairment and/or blindness; ② children with cognitive, hearing, or attention impairments; ③ children wearing myopic corrective glasses or contact lenses; ④ children whose parents did not sign the informed consent paper; and ⑤ children who had received pupil-dilating or pupil-contracting medication within the past week. Since the clinic had ample source of children visiting the clinic during summer vacation, usually at 500-800 children per month (among them about 20% were aged 3-9), we decided to recruited 210 children aged 3-9 to participant in the study.

### 2.3 Tools for VAT

#### The handheld spoon-shaped eye occluder

The handheld spoon-shaped eye occluder, crafted from durable ABS anti-slip plastic, boasts smoothly edges. Measuring 7.63 inches in length and 2.48 inches in width, it comes in a sleek pink or blue color (Figure 2 A).

**Figure 2.**
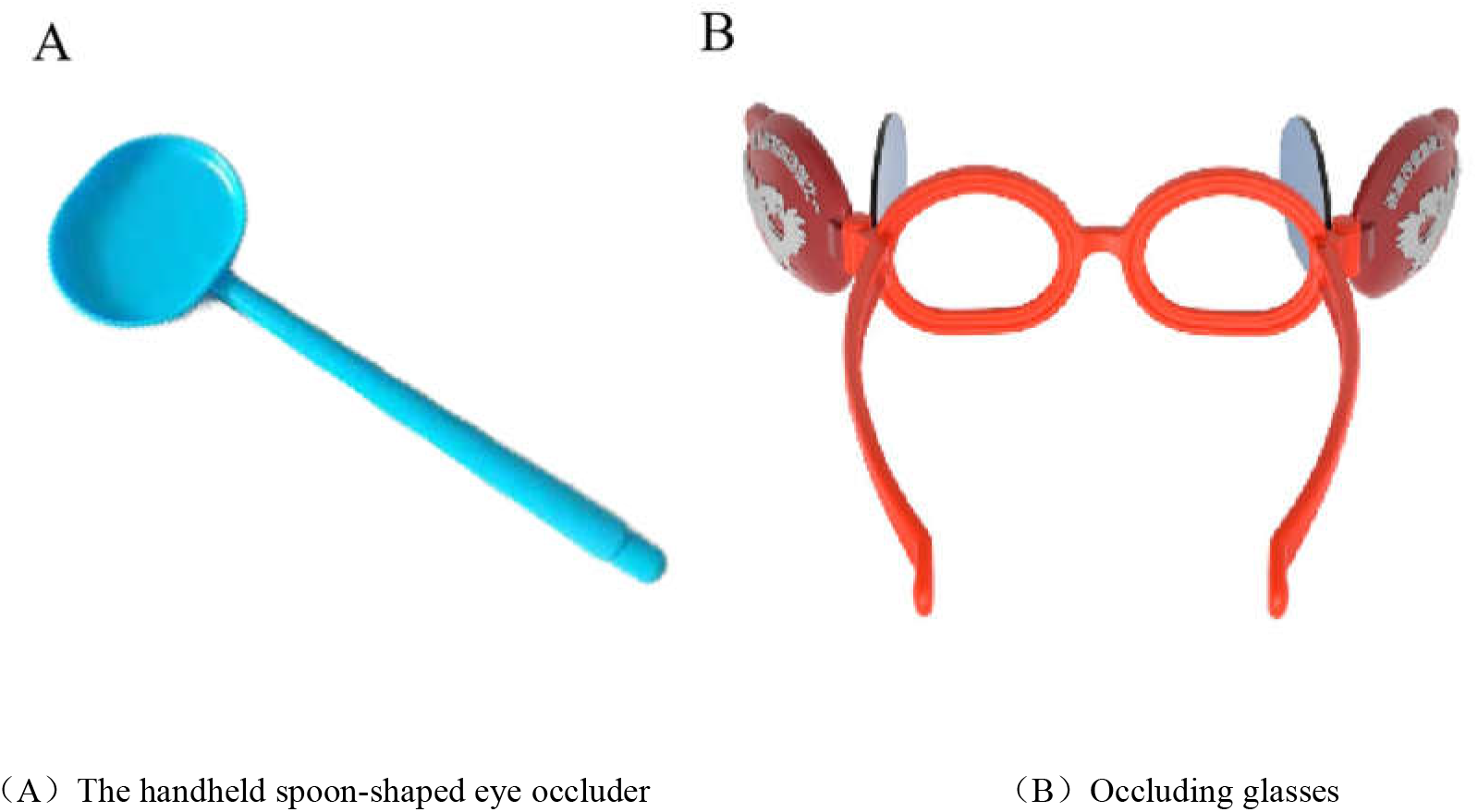
The look of various eye occluders

**Figure 2.**
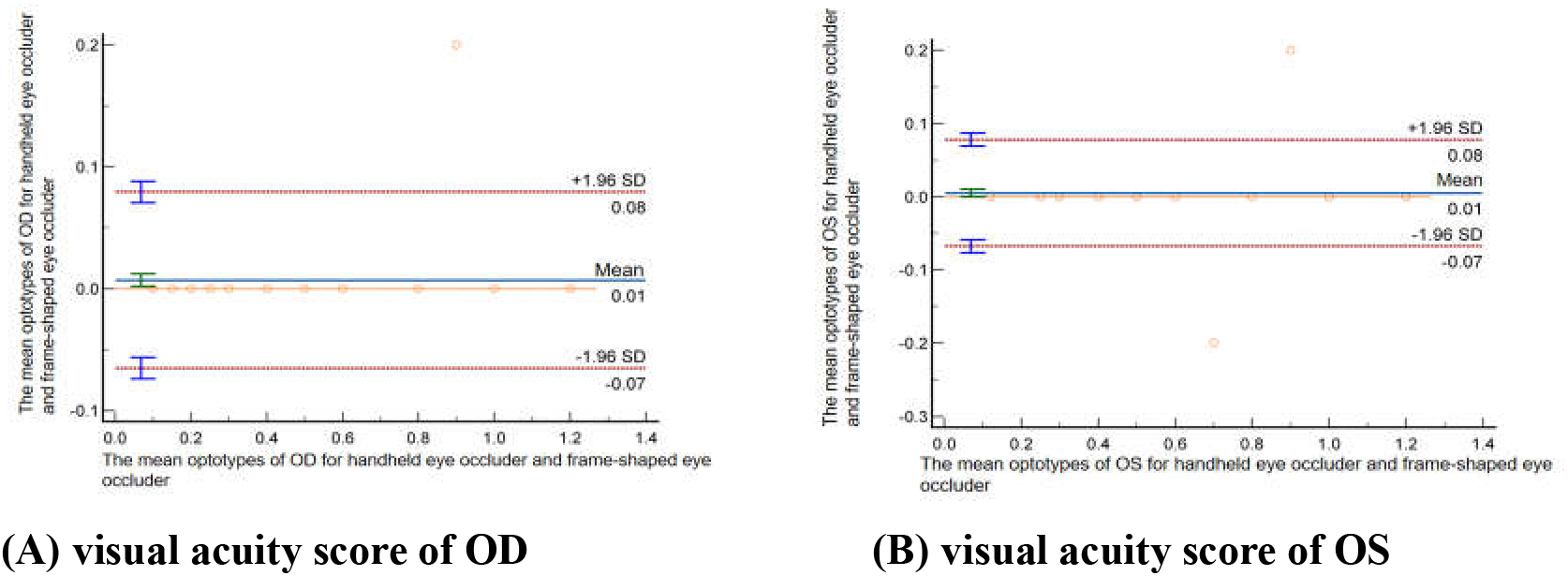
Bland–Altman plots for the Limits of Agreement (LoA) for the paired differences of visual acuity scores measured from the two tools.

#### Occluding glasses

The innovative occluding glasses is a device created by a multidisciplinary team[14], and the design has successfully secured a utility model patent in China (CN202321977842). The tool is composed of one frame, 2 temple pieces, 2 movable blocking lens, and 2 movable patches that had cartoon pictures on the outer most surface. Both the lens and the patches can be extended to the sides and folded toward the bridge of the frame during VAT.Children were asked to put on the glasses and cover one eye with the blocking len and pacth on the same side folded to the bridge, and the blocking len and pacth on the other side extended far wide. Like wise for alternate shielding of the other eye.(Figure 2 B).

### 2.4 Data collection

We utilized a random E chart (2.5 m) to evaluate visual acuity in two batches. Two nurses(One was the observer, the other was the enforcer) performed with a standardized E-shaped optotype chart following Chinese Standard for logarithmic visual acuity charts (GB/T 11533-2011)[15]:①the child was positioned 2.5 meters away from the chart;②proper lighting was ensured for both the room and the E chart;③ the use of the tools: the enforcer had the children cover one eye with a handheld occulder that completely blocked the vision of the covered eye, or had the children put on the occluding glasses and cover one eye with the blocking lense and patch on the same side folded to the bridge;④The enforcer sequentially presented E-optotypes from the top (largest) to bottom (smallest) rows. Participants were instructed to manually indicate the opening direction (up, down, left, or right) of each E-character using a hand gesture or verbal response;⑤Testing progressed row-by-row until the participant failed ≥50% of optotypes within a single row;⑥Switched to the other eye and repeated;⑦recorded visual acuity for each eye by noting the line for which the child correctly identified the orientation.

During the test, the observer nurse stood next to the enforcer nurse and observed how the children behaved, and notched up the moments when they tried to steal a glance from the covered eye.

Upon completion of the 2nd test, the children were invited to evaluate the appearance, convenience, and comfort of both tools through 3 questions: (1) Which eye occluder do you prefer in terms of appearance (the handheld eye occluder or the occluding glasses)? (2) Regarding convenience, which tool do you find more user-friendly? (3) In terms of comfort, which tool do you consider more comfortable? The answer to these questions was to choose between the handheld eye occluder or the occluding glasses.

### 2.5 Quality control

To ensure the objectivity of the study, both the enforcer (Lu Tian) and the observer (Qing Zhou, who is not a member of the research team) were professional ophthalmic nurses. To avoid parental interference with the child, the nurses told the parents beforehand to keep quiet during the VAT of the child. At the end of each day, all the used tools were collected and wiped with disinfecting wipes before they were placed in a UV cabinet. Following 20 minutes of UV radiation, they were ready for later use. Strict inclusion and exclusion criteria were applied at enrollment, and since the vision tests were performed based on physician-ordered prescriptions, nurses took all necessary measures(eg., giving brief rest periods, or allowing the parent to stand beside or behind the child to provide a sense of security. In some cases, parents were also permitted to assist in occluding the non-tested eye, and the nurses strictly instructed them not to cue or prompt the child’s responses) to ensure that uncorrected visual acuity was measured in both eyes for all children. Therefore, no cases were excluded during or after VAT.

### 2.6 Statistic analysis

Excel 2016 and SPSS 26.0 software were utilized for statistical analysis. Descriptive analysis was performed to describe the general characteristics of the study population. Categorical variables were presented as frequencies and proportions, and continuous variables were expressed as mean ± standard deviation if they were normally distrbuted or median and interquartile range[M (P25, P75)],when the quantitative data were skewed. Mann-Whitney U test was used to compare the VAT scores of the two types of eye occluders. The correlation between the outcomes of the two occlusion instruments was assessed using the intra-class correlation coefficient (ICC), and the consistency of the paired visual acuity scores measured by the 2 tools was evaluated through Bland-Altman analysis using the MedCalc software. LoA(limits of agreement) calculated as the mean difference (bias) ± 1.96 times the standard deviation (SD) of the differences between paired measurements, is a statistical tool used to quantify the range within which the differences between two measurement methods are expected to lie for most data points. If the difference between the two measurements falls within the limits of agreement (LoA), it is deemed clinically acceptable, indicating good agreement between the two methods. A P value of <0.05 was considered statistically significant.

## 3. Results

### 3.1 General demographic characteristics (phase 1)

A total of 200 children were enrolled in this study, with a median age of 7 years (interquartile range: 6–8 years). The gender distribution was relatively balanced, with 105 males (52.5%) and 95 females (47.5%). In terms of residence, the sample was predominantly composed of urban children (n = 157, 78.5%), while 43 children (21.5%) were from rural areas. The majority of children (n = 187, 93.5%) had no prior history of vision testing, and only 13 children (6.5%) had previously undergone a vision test. Regarding ocular health, 122 children (61.0%) had normal ocular conditions; 60 children (30.0%) had suspected or confirmed myopia, none of whom were wearing myopia control glasses or contact lenses; and 18 children (9.0%) had strabismus or amblyopia, none of whom were wearing corrective glasses. As shown in Table 1.

**Table 1.**
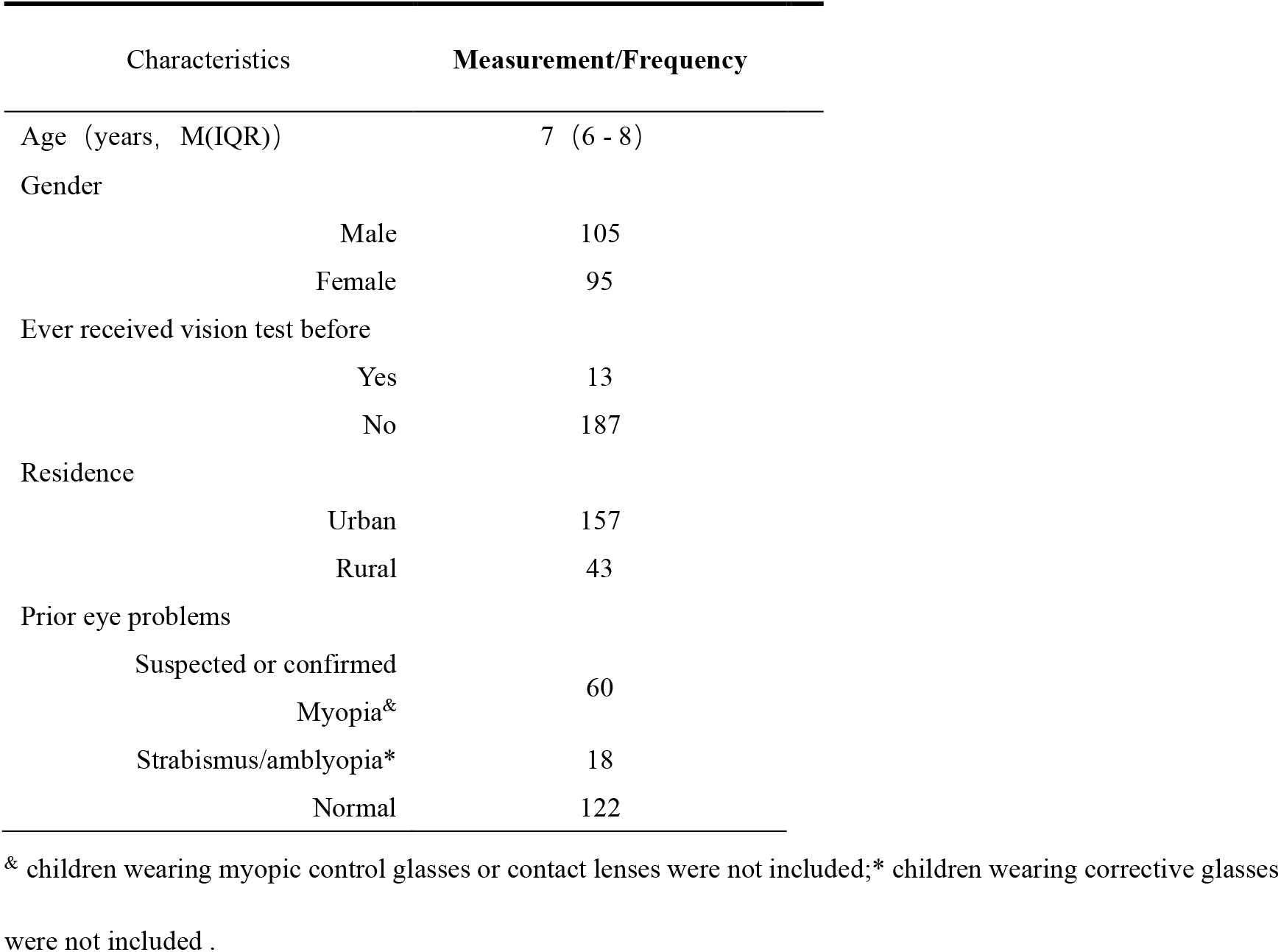
General demographic characteristics of children(n=200)

**Table 2.**
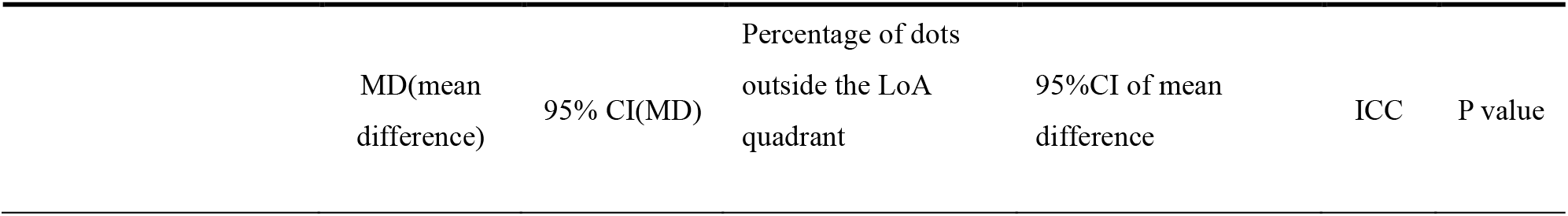
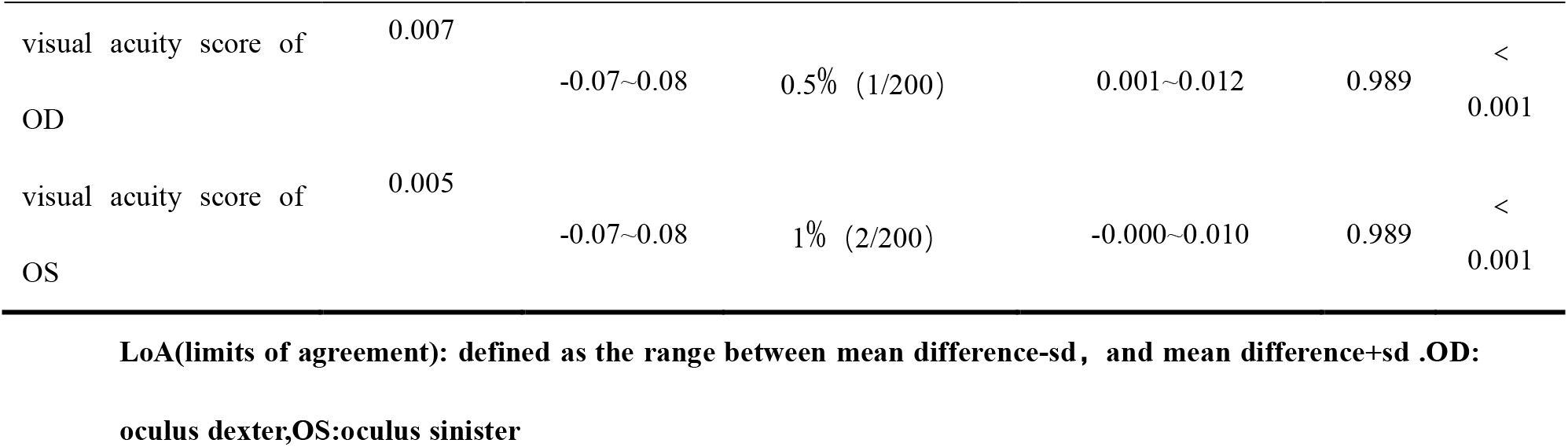
Result of Bland-Altman analysis.

### 3.2 Consistency test of paired visual acuity scores in two tools

The study generated paired visual acuity scores from the two tools. The mean differences in the scores between the two tools were minimal for both eyes. We utilized the visual acuity scores from the handheld spoon-shaped eye occluder as the standard reference values. The ICC values for the paired visual acuity measurements from the two occluders were 0.989 for oculus dexter, likewise for oculus sinister, which exceeds the threshold of 0.7 (P<0.05), suggesting good reliability in paired measurements. In the Bland-Altman plot, the proportion of data points outside the 95% limits of agreement(LoA) was 0.5% (1/200) and 1% (2/200), respectively, confirming that the visual acuity scores from both visual acuity screening tools had a high degree of agreement.

### 3.3 Comparison of results from two tools for visual examination

The median visual acuity score of OD and OS detected in the 200 children using the handheld spoon-shaped eye occluders were both 0.8. Similarly, the median visual acuity score of OD and OS obtained with occluding glasses were also 0.8, with P>0.05 indicating no statistical difference. This finding suggests that the visual acuity results from both tools are consistent, demonstrating no significant difference between them (Table 3).

**Table 3.**
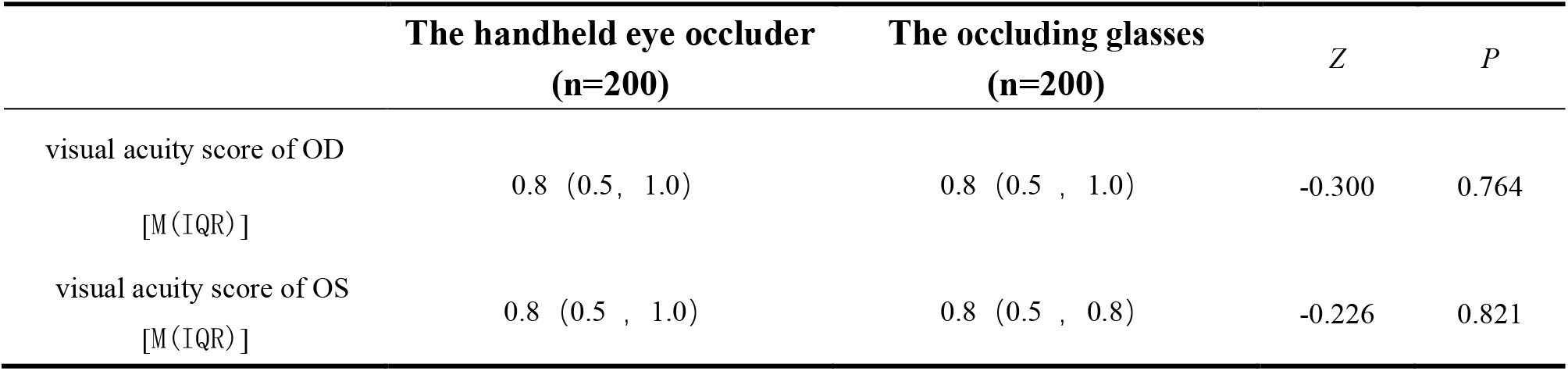
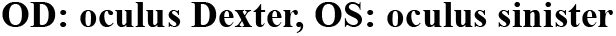
Comparison of results from two types of eyeshield tools.

|  | The handheld eye occluder<br>(n=200) | The occluding glasses<br>(n=200) | Z | P |
| --- | --- | --- | --- | --- |
| visual acuity score of OD<br>[M (IQR)] | 0.8 (0.5, 1.0) | 0.8 (0.5, 1.0) | -0.300 | 0.764 |
| visual acuity score of OS<br>[M (IQR)] | 0.8 (0.5, 1.0) | 0.8 (0.5, 0.8) | -0.226 | 0.821 |

OD: oculus Dexter, OS: oculus sinister

### 3.4 Acceptance of eye covering tools

All children were asked of their acceptance regarding the two eye-covering tools upon completion of the 2nd VAT. Among them, 122 children preferred occluding glasses, while 186 children found handheld spoon-shaped eye occluders to be more comfortable, as presented in Table 5.

**Table 5.**
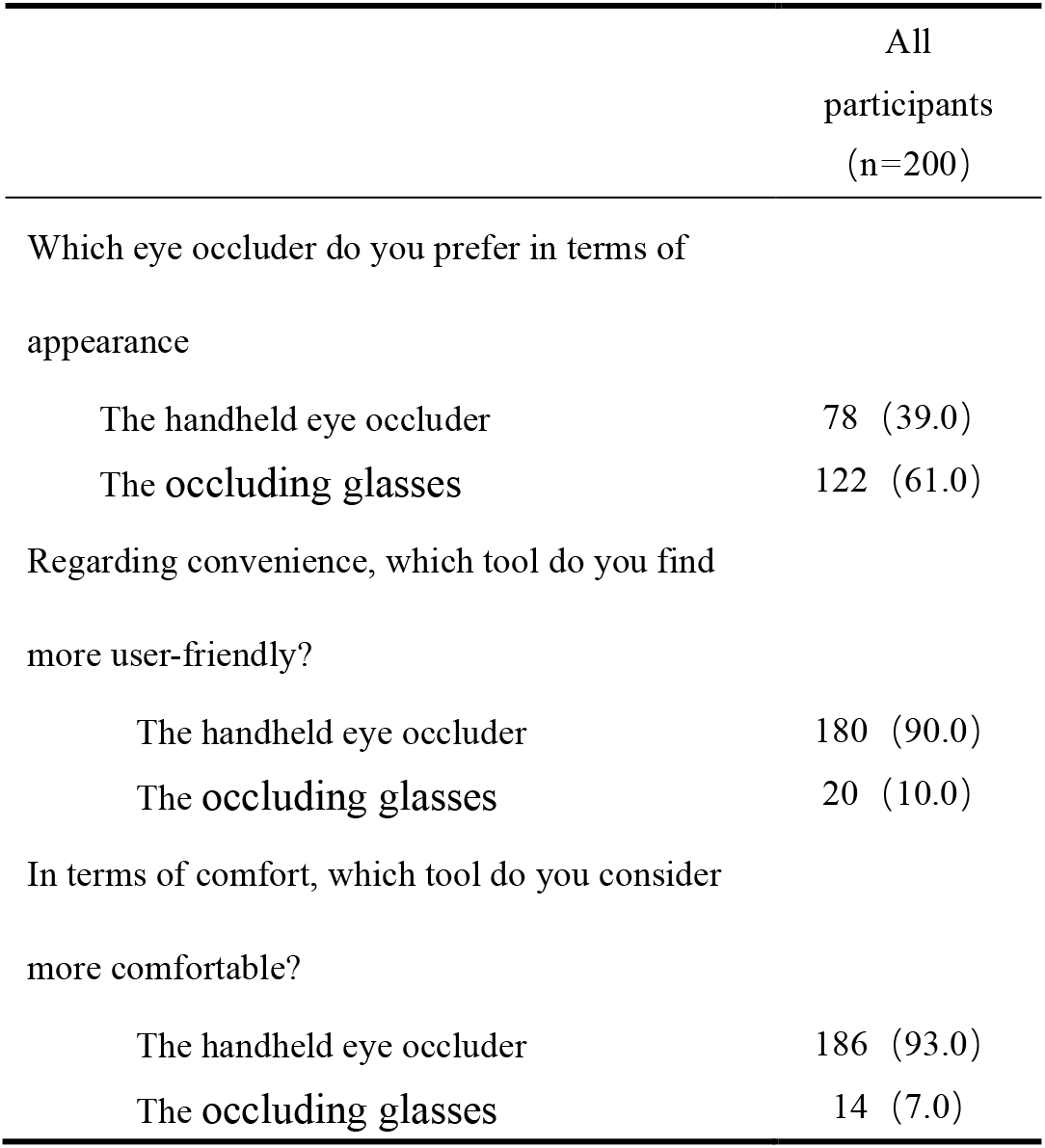
Children’s acceptance of the tools.

## 4. Discussion

In pediatric ophthalmology, the accuracy of a device is a prerequisite for replacing the traditional gold standard. In previous studies, some modified tools improved playfulness but often led to overestimation or underestimation of vision due to light leakage or instability[16, 17]. The study results showed that the mean difference in visual acuity score measured by the 2 tools was close to zero for both eyes, and less than 5% of visual test results were outside LoA (boundary zone). The ICC values for paired measurements generated from 2 tools exceeded 0.7 (P<0.01), indicating good consistency, suggesting that the occluding glasses were on a par with the handheld spoon-shaped eye occluder in terms of test accuracy. It should be emphasized that this study strictly excluded children wearing corrective glasses focusing specifically on assessing uncorrected visual acuity. The Z-test results in this study (Right eye:*Z*= −0.300; Left eye:*Z*= −0.226) further statistically confirm minimal distributional difference between the two measurement methods. The results indicate that in a natural, uncorrected state, the new tool can completely replace the traditional handheld spoon-shaped occluder, justifying its use as a child-friendly tool for VAT in young children. However, since all measurements from the 2 tools were naked vision, the current conclusions cannot be directly extrapolated to children wearing glasses or contact lenses. This suggests that future research is needed to further validate the applicability of this tool in more complex clinical scenarios, such as follow-up examinations involving corrective lenses.

Childhood is a stage in which cognitive abilities and nervous systems are not fully developed[18, 19], during this time, children are particularly susceptible to stress reactions, such as anxiety, feelings of rejection, and fear when faced with unfamiliar situations or environments[20]. Previous studies have shown that when nurses use toys or cartooned tools that are suitable for children’s psychological characteristics, they can effectively lessen children’s resistance during diagnostic tests and treatment process[21,22]. The handheld spoon-shaped eye occluder used in this study represents a purely functional medical device design. Although available in colors such as blue or pink, it lacks any cartoon elements or anthropomorphic features. For children aged 3 to 9 years, this object remains an unfamiliar and boring “tool,” incapable of attracting their attention. The acceptance assessment demonstrated that 61%(122/200) of children preferred the look of the occluding glasses and considered it “more attractive than the handheld one”. However, the traditional occluder still demonstrated a clear advantage in both convenience and comfort (90.0% and 93.0%, respectively). This discrepancy might be attributable to the following factors. First, wearing burden: the traditional occluder is a non-contact handheld device that exerts no physical pressure, whereas glasses-type tools rely on support from the nasal bridge and auricles, which may cause compression discomfort or anxiety about slippage in younger children whose nasal bridges are not yet fully developed. Second, operational complexity: the “pick-up-and-use” nature of the handheld occluder aligns well with children’s short attention spans, while the donning and doffing process of glasses adds both cognitive and motor demands. These findings carry important implications for future product refinement: subsequent development should go beyond playful pattern design and focus more on ergonomic optimization. For instance, lighter materials could be adopted to reduce facial pressure, or addition of fancier elements or popular cartoon figures might help attract children’s attention and reduce discomfort.

## LIMITATIONS

This study has several limitations. As a single-center investigation without prior pilot testing, sample size calculation was not informed by pilot data, which may reduce statistical power. The use of convenience sampling during recruitment may introduce selection bias and limit generalizability. The occlusion glasses feature a fixed-width nose bridge, which cannot fit children with varying facial widths and restricted enrollment to participants aged 3–9 years. Additionally, the device cannot be worn over corrective lenses, so only uncorrected visual acuity was assessed. Its performance for corrected visual acuity testing or when combined with other ophthalmic screening devices remains unevaluated. The limited sample size also prevented age-stratified analyses; further work is needed to examine diagnostic agreement across age strata and different screening settings, such as kindergartens and community screening programs. While measurements obtained with the occlusion glasses agreed well with those from conventional handheld spoon-shaped occluders, children’s acceptance of this novel tool requires validation in interventional implementation studies.

## Data Availability

All data generated or analyzed during this study are included in this published article.

## AI-Assisted Tools Declaration

During the preparation of this manuscript, the authors used Qwen (Tongyi Lab, Alibaba Group, Hangzhou, China), a large language model, for the following purposes: (1) language translation from Chinese to English; (2) language polishing and grammatical refinement; and (3) organization of the writing outline. The authors reviewed and verified all AI-generated content and take full responsibility for the accuracy, integrity, and originality of the final manuscript. No AI tool was involved in the study design, data collection, data analysis, or interpretation of results.

## Author Contributions

### Authors with affiliation 1

**Lu Tian:** Investigation, Methodology (supporting), Resources (supporting), Writing – original draft. **Qing Zhou:** Investigation, Data curation. **Yuhan Song:** Investigation, Data curation. **Meizhen Zhao:** Conceptualization, Resources (supporting), Visualization, Validation, Supervision, Project administration, Writing – review & editing. **Yan Jiang:** Conceptualization, Resources (supporting), Visualization, Validation, Supervision, Project administration.

### Author with affiliation 2

**Shu Yang:** Investigation, Methodology (supporting), Resources (supporting), Writing – original draft.

### Author with affiliation 3

**Yuting Kan:** Writing – review & editing.

All authors have read and agreed to the published version of the manuscript and declare no competing interest.

